# Covid-19: Early evening curfews are not effective and may backfire

**DOI:** 10.1101/2021.04.13.21255091

**Authors:** Sotiris Georganas, Alina Velias, Sotiris Vandoros

## Abstract

**Background:** During the COVID-19 pandemic, some countries have introduced early evening curfews. Several studies try measure the effectiveness of such measures across different countries, but clear identification of effects is elusive.

**Objective:** We examined the impact of an early evening curfew on mobility by studying a shift in curfews from 9pm to 6pm in Greece.

**Data and Methods:** We took advantage of a natural experiment in Greece, where curfews shifted from 9pm to 6pm in one Region, but not in another. We followed a difference-in-differences econometric approach, where we compared trends in mobility in groceries and pharmacies as well as residential spaces before and after the introduction of the 6pm curfew, in the two regions.

**Results:** The relative difference in the time spent in groceries and pharmacies between the two regions before and after the intervention, is statistically insignificant [coeff: −9.95; 95%CI −44.358 to 24.458]. The relative increase in time spent at residential spaces after the 6pm curfew was only 4.625 percentage points [coeff: 4.625; 95%CI 1.412 to 7.838].

**Conclusions:** We found that the 6pm instead of 9pm curfew in Athens led to a 4.63 percentage point relative increase in time spent at home and had no effect on time spent in groceries and pharmacies. Considering that this was a result of a 18.75% reduction in hours where people were allowed to leave home, it seems that the early evening curfew led to more crowding in indoor spaces – which may facilitate the spread of disease. Interventions should be based on a thorough analysis of human behaviour, that anticipates substitution of activities.

## 1. Background

There is an ongoing debate about non-pharmaceutical interventions and how effective they are in tackling the Covid-19 pandemic.^1^ Such measures are most often introduced jointly, so disentangling the effects of individual measures is challenging, while finding appropriate control groups is not always straightforward.

An increasing number of studies suggest that stringency of the lockdowns measures does not make a difference in infection prevalence or related deaths.^2^ Of course, such studies are often challenged by measurement and identification issues. Also, there are examples of evidence that the ‘signal (inducing voluntary behavior changes) is important in contrast to the actual regulation (mandated behavior changes)’.^3^ In general though, the economic literature, as well as legal research, suggest that restricting one human activity often leads to substitution by others, as humans seek alternatives. And there is a strictness level beyond which extra measures can actually backfire. In this paper we show that early curfews may be one of these cases, where excessive strictness can lead to the opposite of the intended result.

Why would reducing the time window during which people are allowed to leave their homes either fail to achieve the desired greater reduction in virus spread or, even, backfire by contributing towards the spread? One straight-forward reason is that people do not fully reduce the activity proportionately to the strictness of measures – they reallocate part of it towards options that are still allowed. For example, mobile tracker data in the US shows a large reallocation of consumer activity from “nonessential” to “essential” businesses as well as from restaurants and bars toward groceries and other food sellers.^4^ Whilst these studies do not measure the resulting congestion in the essential businesses, we argue that it is likely to be high, resulting in greater risk of virus spread.

In our study, we took advantage of within-country heterogeneity in the timing of the introduction of an early evening curfew to evaluate this measure in tackling the Covid-19 pandemic. The advantage of our data is that we can cleanly identify the effect using a sudden and singular change in the lockdown rules.

While cross country studies are definitely also very useful, they suffer from several drawbacks absent in our method. On the one hand countries differ in important characteristics that affect the performance of measures (availability of ICUs, the state of the health system), but crucially they also differ in the way they measure the pandemic itself. Recorded cases are biased and since testing methods are not homogeneous across countries, the bias is heterogeneous, differing greatly across countries.^5^ On the other hand, the measures are not the same across countries, and seldom are enacted alone within a specific country. Usually, a complicated bundle of measures is enacted on the same day (and some measures almost always come together, such as closing several levels of schooling along with other face to face activities) which greatly complicates isolating the effect of a single measure. In this paper we use a natural experiment in a single European country to clearly identify the effect of early curfews, using a difference in difference approach, comparing a region affected by the curfew to neighbouring regions.

## 2. Data and Methods

While a 9pm-5am curfew applied in Greece since November 2020, a 6pm-5am *weekend* curfew was introduced on 6 February 2021 in the Attica region (which includes the capital city of Athens) as a response to increasing Covid-19 cases. We studied the impact of the 6pm curfew on human activity using mobility data from Google Community Mobility Reports^6^. In particular, we focused on time spent at home, and time spent at groceries/pharmacies. Staying at home is considered a goal of lockdown measures, to limit the spread of the novel coronavirus; while it has been shown that indoor spaces such as supermarkets may facilitate transmission.^7^ Google mobility reports show how time spent in different places compares to the baseline, which was the “median value for the corresponding day of the week, during the 5-week period Jan 3 – Feb 6, 2020”,^6^ before the pandemic started in Greece.^a^

A simple before-after analysis to evaluate the impact of the policy on mobility may not be reliable as other factors affecting mobility may change (e.g. the weather), which is why we used the Epirus & Western Macedonia Region (where the curfew remained at 9pm during the study period) as a control group. We studied the difference in the differences in mobility between these two areas in the five weekends before and the four weekends after the introduction of the 6pm curfew in Attica using a difference-in-differences (DiD) ordinary least squares econometric estimator. A DiD model compares trends in the outcome in the treatment group and a control group before and after a particular intervention, which is used extensively in the literature for causal inference.^8-9^ In such empirical models, there is a treatment group dummy, which takes the value of 1 for the group that underwent a treatment; and a treatment period dummy, which takes the value of 1 for the time after the intervention. The interaction of the treatment group dummy and the treatment period dummy gives us the main variable of interest.

In the DiD model, the dependent variable is the percentage change in time spent in a particular type of location compared to the baseline. In one model we study groceries and pharmacies, and in the other we study residential spaces. We included a dummy variable for the Attica region which is the treatment group (1 for observations on Attica and 0 for Epirus & Western Macedonia – the control group), and a dummy variable which takes the value of one in the post-treatment period (from 6 February onwards) and zero otherwise. The interaction between the two shows whether the intervention had an effect on relative trends. We used robust standard errors in regressions.

Figure 1 shows the trends in mobility the two regions before and after the 6pm curfew intervention (in week 11). The results of the DiD regressions are presented in Table 1. When considering the effect on time spent in groceries and pharmacies (column 1), the coefficient of the DiD interaction term, that shows the difference in the difference between the two regions, is statistically insignificant [coeff: −9.95; 95%CI −44.358 to 24.458]. This suggests that there was no change in the relative trends in visits to groceries and pharmacies in Attica compared to the control group after the intervention. Column 2 shows the results of the model with time spent at residential spaces as outcome. The DiD interaction term is positive and statistically significant [coeff: 4.625; 95%CI 1.412 to 7.838], suggesting that the relative increase in time spent at residential spaces after the 6pm curfew was only 4.625 percentage points.

**Table 1.** Results of the Difference-in-Differences regressions

|  | grocery & pharmacy | residential |
| --- | --- | --- |
| DiD interaction term (Attica*week 6 onwards) | -9.95<br>[-44.358 - 24.458] | 4.625***<br>[1.412 - 7.838] |
| Week 6 onwards (treatment period) | 1.725<br>[-23.336 - 26.786] | -4.2***<br>[-6.668 - -1.732] |
| Attica dummy variable (treatment group) | -2.3<br>-66.904 | 1<br>[-1.086 - 3.086] |
| Constant term | 3.9<br>[-20.211 - 28.011] | 11.2***<br>[9.948 - 12.452] |
| Observations | 36 | 36 |
| R-squared | 0.0282 | 0.4825 |
| F-statistic | 3.37 | 7.71 |

The dependent variable is the change in time spent in the two types of locations compared to the baseline. Robust 95% confidence intervals in brackets. \*\*\* p<0.01, \*\* p<0.05, \* p<0.1.

**Figure 1.**
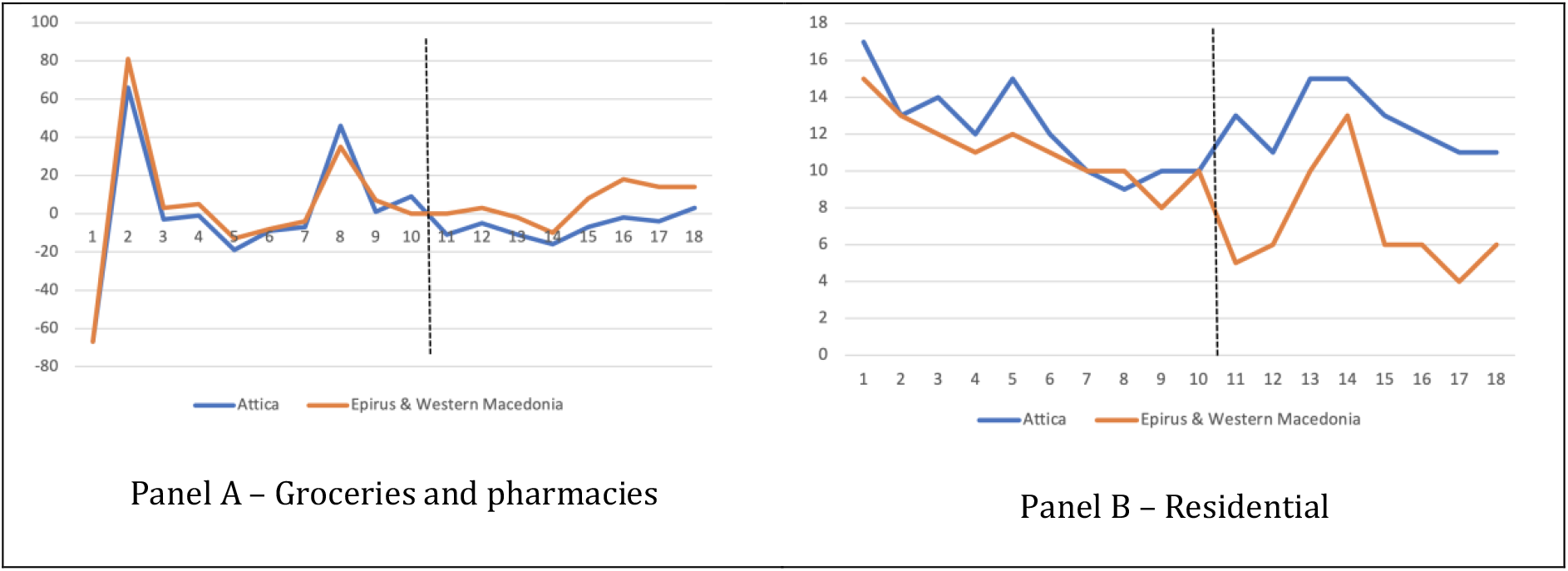
Trends in mobility before and after the curfew. Vertical line shows the introduction of the 6pm curfew in Attica

The results of the econometric analysis show that a reduction in the time when people were allowed to go outside by 3 hours (a 18.75% decrease) led to a 4.63 percentage point increase in time spent at home and had no effect on time spent in groceries/pharmacies, in relative terms. As the change in activities is much smaller than the change in time, the early curfew led to greater crowding. Especially with regards to grocery stores, the same amount of time appears to be spent in a narrower time window. As more people were present simultaneously in high-risk places such as supermarkets, the early curfew backfired, possibly leading to greater disease transmission. Finding the exact impact on crowding is not straightforward, as Google mobility data do not show what time of the day these activities took place, or the density of activities during the day.

## 4. Conclusions

We found that the 6pm instead of 9pm curfew in Athens led to a 4.63 percentage point relative increase in time spent at home and had no effect on time spent in groceries and pharmacies. Considering that this was a result of a 18.75% reduction in hours where people were allowed to leave home, it seems that the early evening curfew led to more crowding in indoor spaces – which may facilitate the spread of disease. Our findings add to existing evidence from Toulouse,^10^ that suggests that a 6pm curfew backfired.

This study is subject to limitations. The outcome is mobility rather than infections. Although certain environments such as supermarkets have a higher likelihood of spreading the disease,^7^ our data do not show the actual impact on Covid-19. However, such an effect would be extremely challenging to disentangle, even with clinical data for the following reasons: (a) Other factors such as variants that may be more transmissible may apply, distorting the effect on actual health outcomes; (b) there is dispersion in the time lag between infection and symptoms or hospitalisation or death; and (c) the effect might show via second-hand transmission. For example, individuals who first contract SARS-CoV-2 due to more crowding may be younger people who are often asymptomatic,^11^ and may pass the virus on to others with a longer lag. Furthermore, Google mobility data do not include a breakdown by time of day, so we cannot be sure how the distribution of mobility changes, and the extent to which crowding occurs. While accurately estimating the effect is not possible, the data show a clear direction towards more crowding in certain spaces.

Overall, lockdowns and other measures are needed to tackle Covid-19. However, it is important to design smart measures that do not lead to substitution by activities that contribute further to spreading the virus. It seems that in some measures can indeed be *too* strict, even if containing the disease is the only goal.

## Data Availability

The data are publicly available already,

## Authors’ contributions

All authors contributed equally to the study.

## Ethics

We did not use any individual-level data, so ethics approval was not required.

## Data availability

Data were obtained from Google community mobility reports and are publicly available online

## Footnotes

a Apple mobility trends reports are also available. However, these only cover driving, walking and commuting, and captures the volume of requests, rather than actual mobility. Furthermore, Apple makes data available for Attica and Greece, so the control group would be contaminated by areas where the treatment applies. Despite these issues, any evidence we could get out of the Apple data points at exactly the same direction as the results using the Google data.

